# Recovery of inhibitory control prefrontal cortex function in inpatients with heroin use disorder: a 15-week longitudinal fMRI study

**DOI:** 10.1101/2023.03.28.23287864

**Authors:** Ahmet O. Ceceli, Yuefeng Huang, Pierre-Olivier Gaudreault, Natalie E. McClain, Sarah G. King, Greg Kronberg, Amelia Brackett, Gabriela N. Hoberman, John H. Gray, Eric L. Garland, Nelly Alia-Klein, Rita Z. Goldstein

## Abstract

**Importance:** Heroin addiction and related mortality impose a devastating toll on society, with little known about the neurobiology of this disease or its treatment. Poor inhibitory control is a common manifestation of prefrontal cortex (PFC) impairments in addiction, and its potential recovery following treatment is largely unknown in heroin (or any drug) addiction.

**Objective:** To study inhibitory control brain activity in iHUD and HC, before and after 15 weeks of inpatient treatment in the former.

**Design:** A longitudinal cohort study (11/2020-03/2022) where iHUD and HC underwent baseline and follow-up fMRI scans. Average follow-up duration: 15 weeks.

**Setting:** The iHUD and HC were recruited from treatment facilities and surrounding neighborhoods, respectively.

**Participants:** Twenty-six iHUD [40.6±10.1 years; 7 (29.2%) women] and 24 age-/sex-matched HC [41.1±9.9 years; 9 (37.5%) women].

**Intervention:** Following the baseline scan, inpatient iHUD continued to participate in a medically-assisted program for an average of 15 weeks (abstinence increased from an initial 183±236 days by 65±82 days). The HC were scanned at similar time intervals.

**Main Outcomes and Measures:** Behavioral performance as measured by the stop-signal response time (SSRT), target detection sensitivity (*d’*, proportion of hits in go vs. false-alarms in stop trials), and brain activity (blood-oxygen level dependent signal differences) during successful vs. failed stops in the stop signal task.

**Results:** As we previously reported, at time 1 and as compared to HC, iHUD exhibited similar SSRT but impaired *d’* [*t*(38.7)=2.37, *p*=.023], and lower anterior and dorsolateral PFC (aPFC, dlPFC) activity (*p*<.001). Importantly, at time 2, there were significant gains in aPFC and dlPFC activity in the iHUD (group*session interaction, *p*=.002); the former significantly correlated with increases in *d’* specifically in iHUD (*p*=.012).

**Conclusions and Relevance:** Compared to HC, the aPFC and dlPFC impairments in the iHUD at time 1 were normalized at time 2, which was associated with individual differences in improvements in target detection sensitivity. For the first time in any drug addiction, these results indicate a treatment-mediated inhibitory control brain activity recovery. These neurobehavioral results highlight the aPFC and dlPFC as targets for intervention with a potential to enhance self-control recovery in heroin addiction.

**Key points:** *Question:* Does inhibitory control brain function, a common impairment in drug addiction, recover with treatment in inpatient individuals with heroin use disorder (iHUD)?

*Findings:* In this longitudinal cohort study, 26 inpatient iHUD and 24 healthy controls (HC) performed a stop-signal task during functional MRI twice, separated by an average of 15 weeks. In the iHUD, we found lower anterior and dorsolateral prefrontal cortex (comprising the cognitive control network) signaling at baseline, which increased after 15 weeks of treatment, as associated with behavioral improvements.

*Meaning:* As the opioid epidemic continues, our results indicate potential therapeutic targets to enhance neural function underlying self-control in heroin addiction.

## Introduction

The opioid epidemic continues to exert its devastating toll with over 100,000 overdose-related deaths in 2021 alone ^1^, rendering paramount the need for effective treatments. However, the neurobehavioral mechanisms underlying recovery of symptoms associated with opioid (e.g., heroin) addiction remain unclear. Given its putative contribution to drug use and relapse in drug addiction, deficient inhibitory control may be an important symptomatic target ^2,3^. This core symptom of drug addiction is associated with downregulation of prefrontal cortex (PFC) regions (see ^4–6^ for reviews) comprising the cognitive control network, including the inferior frontal gyrus (IFG), supplementary motor area, and dorsolateral PFC (dlPFC) ^7^. Recently, using the hallmark measure of inhibitory control, the stop-signal task (SST), we reported lower inhibitory control performance (as indicated in *d*’, or target detection sensitivity) and anterior PFC (aPFC) and dlPFC activity in 41 abstinent individuals with heroin use disorder (iHUD) as compared to 24 demographically-matched healthy controls (HC) ^8^. While this cross-sectional neuroimaging study implicated the PFC in inhibitory control deficits in heroin addiction, exploring whether treatment/abstinence normalizes these neurobehavioral impairments requires longitudinal (within-subject) efforts.

Although an intuitive notion, the empirical evidence for the potential recovery of inhibitory control-related neural function in drug addiction is scarce. A recent review reported very few longitudinal fMRI studies of inhibitory control-related processes in individuals with substance use disorders ^9^. In a series of three clinical trials, 44 individuals with cocaine use disorder performed the color-word Stroop fMRI task at two time points, separated by 8 or 12 weeks of cognitive behavioral and/or pharmacological intervention. There were no significant changes in cognitive control network engagement at follow-up compared to baseline ^10^ (see ^11–13^ for the smaller individual preliminary trials). Using the addiction Stroop fMRI task (a drug-word modification of the classic color-word Stroop task; reviewed in ^14^) at two time points, separated by six months of abstinence, we showed increased BOLD signal in the dopaminergic midbrain in 15 individuals with cocaine use disorder. This increase in midbrain activity correlated with lower simulated drug seeking, indicative of enhanced self-control (or lower drug-biased salience attribution) in a drug context ^15^. Specifically in heroin addiction, a single study examined longitudinal changes in brain activity during a Go/No-Go task. Twenty-one methadone-maintained iHUD (abstinent for at least 3 months) had increased right IFG function after a year of treatment ^16^, suggesting that PFC activity may serve as a sensitive marker of recovery in treatment. This study did not include a HC group and hence the potential confounds of time (and task repetition) were not accounted.

To summarize, only a handful of fMRI longitudinal studies targeted inhibitory control-related recovery with time (as a function of abstinence and/or treatment) in individuals with substance use disorder, and a lower proportion included a HC group. Further, unlike the SST, which estimates the ability to stop after an already initiated response—a defining component of inhibitory control ^17^, these studies used Stroop and Go/No-Go tasks, the former estimating attentional interference ^18^ and the latter capturing response selection ^19^. To the best of our knowledge, the potential recovery of inhibitory control brain function in heroin (or any drug) addiction remains untested. Therefore, we employed a well-validated SST during fMRI in a longitudinal design to inspect the potential time-dependent recovery of the impaired neural processes underlying inhibitory control in inpatient iHUD as compared to demographically-matched HC. We hypothesized, in iHUD, 1) impaired inhibitory control behavioral performance and PFC activity at baseline compared to HC, 2) significant recovery of PFC activity after 15 weeks of treatment, and 3) PFC inhibitory control recovery from baseline to follow-up to be associated with improvements in behavioral performance.

## Materials and Methods

### Participants

Twenty-six iHUD (mean age=40.6±10.1 years; 7 women) were recruited from a medication-assisted inpatient rehabilitation facility and 24 age- and sex-matched HC (mean age=41.1±9.9 years; 9 women) were recruited from the surrounding community for matching purposes through advertisements and word of mouth. See Table 1 for sample descriptive statistics and the supplement for exclusion criteria. Twenty-five iHUD and 22 HC in the current study were part of the previous study sample in our recent cross-sectional report that included 41 iHUD and 24 HC ^8^. The Icahn School of Medicine at Mount Sinai’s institutional review board approved study procedures, and all participants provided written informed consent.

**Table 1.**
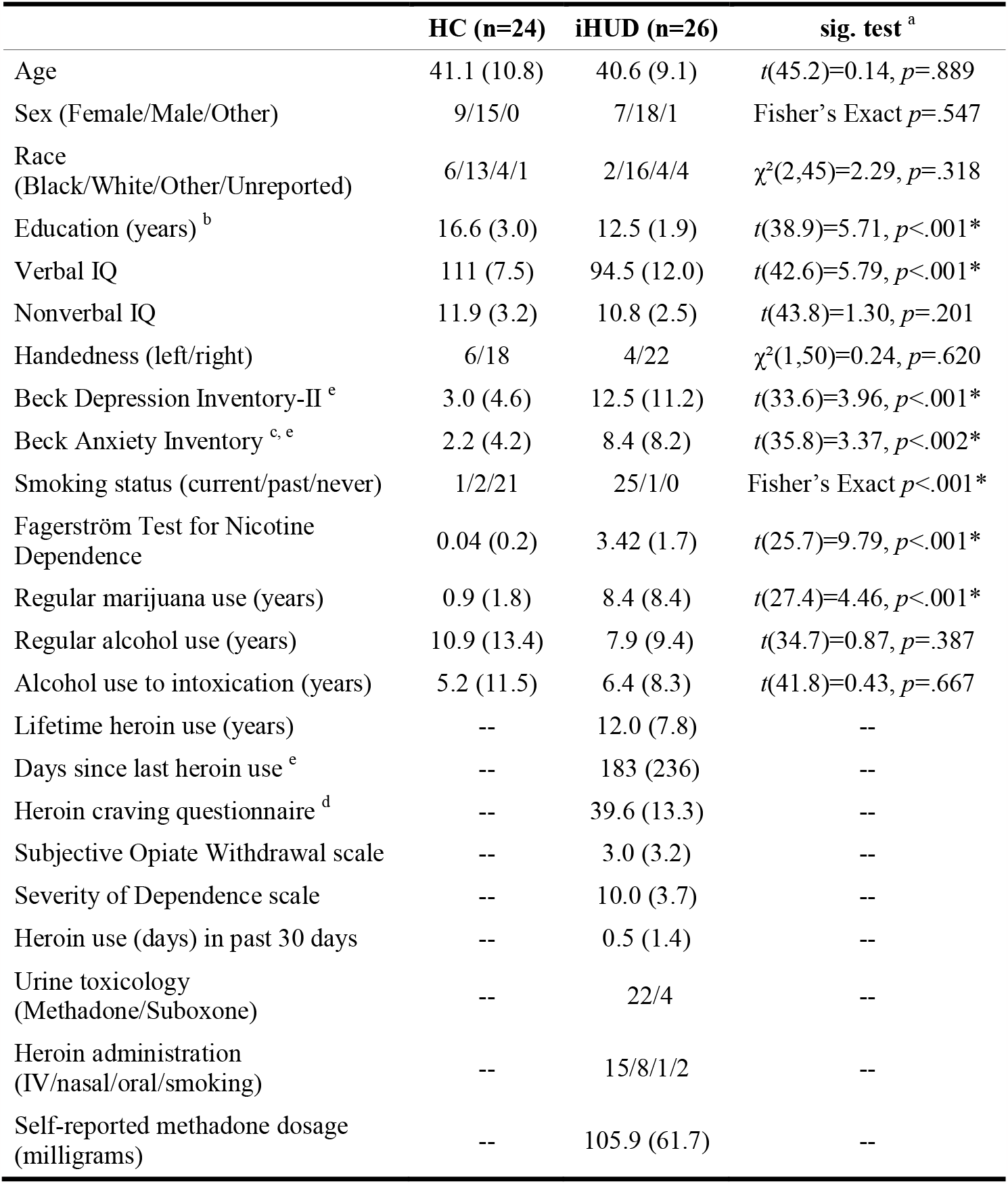
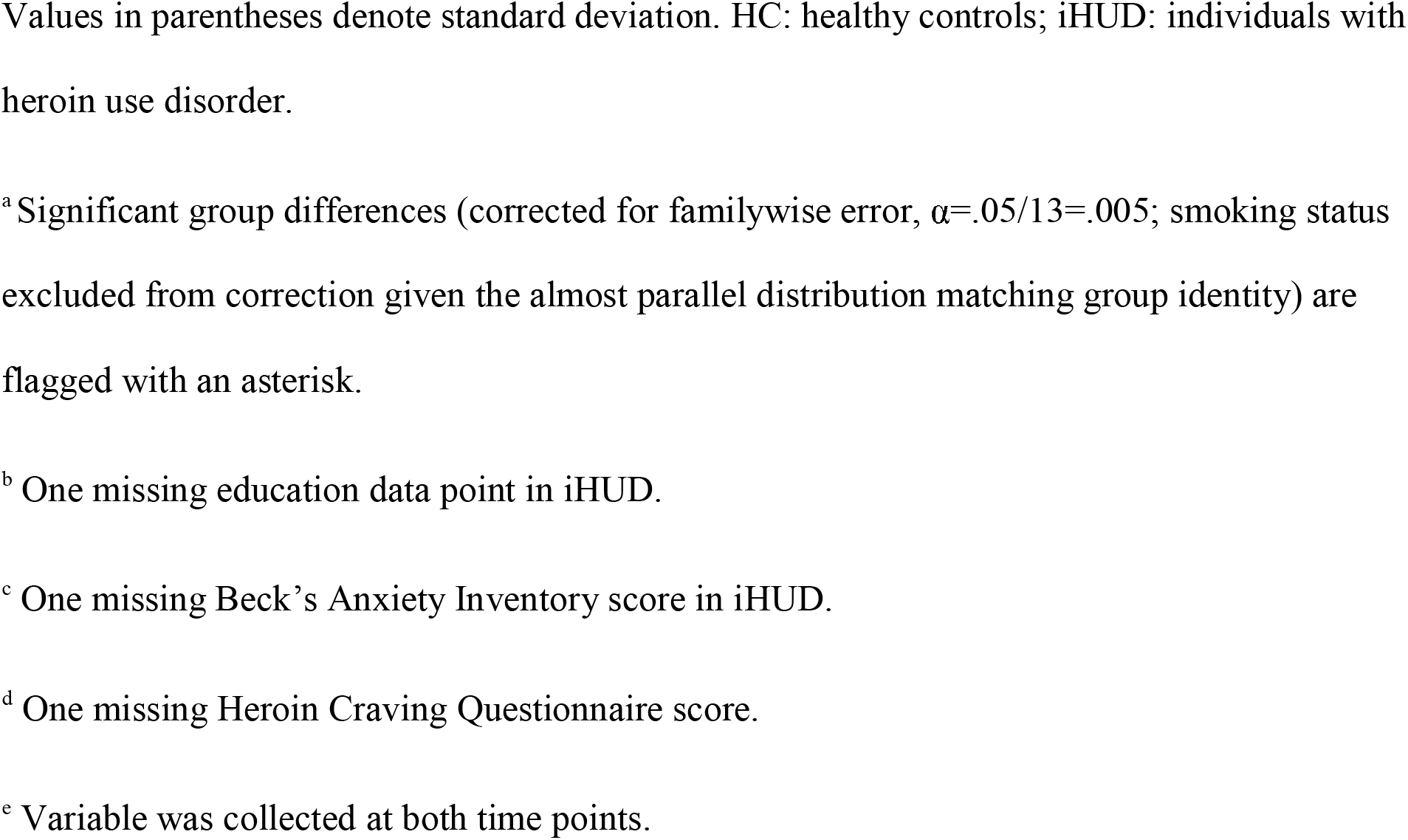
Sample profile.

|  | HC (n=24) | iHUD (n=26) | sig. test <sup>a</sup> |
| --- | --- | --- | --- |
| Age | 41.1 (10.8) | 40.6 (9.1) | $t(45.2)=0.14, p=.889$ |
| Sex (Female/Male/Other) | 9/15/0 | 7/18/1 | Fisher's Exact $p=.547$ |
| Race<br>(Black/White/Other/Unreported) | 6/13/4/1 | 2/16/4/4 | $\chi^2(2,45)=2.29, p=.318$ |
| Education (years) <sup>b</sup> | 16.6 (3.0) | 12.5 (1.9) | $t(38.9)=5.71, p<.001^*$ |
| Verbal IQ | 111 (7.5) | 94.5 (12.0) | $t(42.6)=5.79, p<.001^*$ |
| Nonverbal IQ | 11.9 (3.2) | 10.8 (2.5) | $t(43.8)=1.30, p=.201$ |
| Handedness (left/right) | 6/18 | 4/22 | $\chi^2(1,50)=0.24, p=.620$ |
| Beck Depression Inventory-II <sup>e</sup> | 3.0 (4.6) | 12.5 (11.2) | $t(33.6)=3.96, p<.001^*$ |
| Beck Anxiety Inventory <sup>c, e</sup> | 2.2 (4.2) | 8.4 (8.2) | $t(35.8)=3.37, p<.002^*$ |
| Smoking status (current/past/never) | 1/2/21 | 25/1/0 | Fisher's Exact $p<.001^*$ |
| Fagerström Test for Nicotine<br>Dependence | 0.04 (0.2) | 3.42 (1.7) | $t(25.7)=9.79, p<.001^*$ |
| Regular marijuana use (years) | 0.9 (1.8) | 8.4 (8.4) | $t(27.4)=4.46, p<.001^*$ |
| Regular alcohol use (years) | 10.9 (13.4) | 7.9 (9.4) | $t(34.7)=0.87, p=.387$ |
| Alcohol use to intoxication (years) | 5.2 (11.5) | 6.4 (8.3) | $t(41.8)=0.43, p=.667$ |
| Lifetime heroin use (years) | -- | 12.0 (7.8) | -- |
| Days since last heroin use <sup>e</sup> | -- | 183 (236) | -- |
| Heroin craving questionnaire <sup>d</sup> | -- | 39.6 (13.3) | -- |
| Subjective Opiate Withdrawal scale | -- | 3.0 (3.2) | -- |
| Severity of Dependence scale | -- | 10.0 (3.7) | -- |
| Heroin use (days) in past 30 days | -- | 0.5 (1.4) | -- |
| Urine toxicology<br>(Methadone/Suboxone) | -- | 22/4 | -- |
| Heroin administration<br>(IV/nasal/oral/smoking) | -- | 15/8/1/2 | -- |
| Self-reported methadone dosage<br>(milligrams) | -- | 105.9 (61.7) | -- |

Values in parentheses denote standard deviation. HC: healthy controls; iHUD: individuals with heroin use disorder.
<sup>a</sup> Significant group differences (corrected for familywise error, $\alpha=.05/13=.005$ ; smoking status excluded from correction given the almost parallel distribution matching group identity) are flagged with an asterisk.
<sup>b</sup> One missing education data point in iHUD.
<sup>c</sup> One missing Beck's Anxiety Inventory score in iHUD.
<sup>d</sup> One missing Heroin Craving Questionnaire score.
<sup>e</sup> Variable was collected at both time points.

### Diagnostic and Clinical Procedures

All iHUD were inpatients in a facility where they attended courses/treatments including relapse prevention, Seeking Safety therapy (a present-focused counseling model to help people attain safety from trauma and/or substance abuse), and anger management. A comprehensive clinical diagnostic and substance use interview was conducted at baseline, consisting of the Mini International Neuropsychiatric Interview 7^th^ edition ^20^ and the Addiction Severity Index 5^th^ edition ^21^. Drug dependence severity, craving, and withdrawal symptoms were assessed using the Severity of Dependence Scale ^22^, the Heroin Craving Questionnaire [a modified version of the Cocaine Craving Questionnaire ^23^] and the Subjective Opiate Withdrawal Scale ^24^, respectively.

Nicotine dependence severity was measured using the Fagerström Test for Nicotine Dependence ^25^. All iHUD met DSM-5 criteria for opioid use disorder (with heroin as the primary drug of choice/reason for treatment). Substance use-related psychiatric comorbidities commonly observed in individuals with drug addiction (27, 28) were either in partial or sustained remission at baseline with no current comorbidities found in the HC (see supplement). All iHUD were abstinent (for an average of 183 days during their first study session) and under medication-assisted treatment (confirmed via toxicology on both sessions and dosage in mg collected via self-report at baseline). Beck’s Depression and Anxiety Inventories, cue-induced drug craving (via self-reported ratings to picture stimuli from an in-house drug cue reactivity task ^26^) and number of days since last heroin use were collected at both sessions. Table 1 details heroin route of administration, nicotine/alcohol/cannabis use, urine toxicology, and medication dose, in addition to demographics, neuropsychological and additional drug use measures.

Following baseline assessments including MRI (see below), iHUD were randomly assigned to one of two types of group therapy (i.e., Mindfulness Oriented Recovery Enhancement or support group) consisting of weekly two-hour sessions led by trained therapists. Data from participants in both treatment groups were combined for the present analyses to assess the general treatment effect. Therapy-specific results of the clinical trial associated with this study (NCT04112186) will be reported separately (see supplement for therapy group details).

### Magnetic Resonance Imaging

Participants underwent two MRI scans (baseline and follow-up) separated by an average of 106.0±34.3 (range=62-185) days in iHUD and 93.7±50.6 (range=56-281) days in HC, with no significant group differences in days between scans [*t*(40)=1.00, *p*=.321]. Scanning was conducted on a Siemens 3T Skyra (Siemens Healthcare, Erlangen, Germany), using a 32-channel head coil (see supplement for scanning and preprocessing parameters).

### Experimental Paradigm

#### The Stop-Signal Task

In agreement with the latest guidelines in estimating inhibitory control ^27^, we administered the recently revised SST, *STOP-IT* ^28^, modified for the fMRI context and presented via jsPsych ^29^ as recently reported ^8^. Participants made index or middle finger button presses using an MR-compatible response glove that corresponded to the left or right white arrows (or the opposite in left-handed participants), respectively, as quickly and accurately as possible. In 25% of these go trials, the white arrow (the go signal) changed to red (the stop-signal) after a variable delay (the stop-signal delay, or SSD), to which participants were instructed to stop their response. The SSD was set to an initial duration of 200 ms and tracked the participant’s stopping ability, increasing by 50 ms after successful stops (making the next stop trial more difficult), and decreasing by 50 ms after failed stops (making the next trial easier). The task was administered over two fMRI runs, separated by a brief interval (10 s) when we displayed interim average response time (RT) in ms, proportion of missed go trials, and proportion of correct stops to enhance task compliance.

### Behavioral Data Analyses

We followed well-established parameters in line with the horse-race model of inhibitory control ^30^ to estimate stop-signal RT (SSRT; lower values indicate quicker performance), using the package *ANALYZE-IT* via the integration method (see ^27^ for details). We further inspected *d’*, derived from a Z-transformation of hit (proportion of correct responses to go trials) and false alarm (proportion of responses following a stop-signal) rates, with higher *d’* values reflecting higher sensitivity in detecting targets over non-targets.

Given our prior cross-sectional report of SST impairment in iHUD ^8^, and our current *a priori* interest in treatment-related effects, here we focused on performance differences between sessions, with cross-sectional baseline and follow-up behavioral results reported for completeness. We used linear mixed models [group (iHUD, HC) and session (baseline, follow-up) as fixed factors, subject as random factors] to inspect group differences in SSRT and *d’* change (Δ), corrected for familywise error for these two measures of interest (α=.05/2=.025). We further tested for contributions to SSRT and *d’*, as well as their Δ between sessions, of potentially explanatory demographic and neuropsychological measures that differed between groups and/or sessions. Within iHUD, we similarly examined the potential relationships between SSRT/*d*’ and baseline heroin use-related Table 1 measures including methadone dosage (used by most iHUD), as well as commonly used drugs (alcohol, nicotine, marijuana) that indicated group differences (see supplement).

### BOLD-fMRI Data Analyses

*Go Success, Go Fail, Stop Success*, and *Stop Fail* events were sampled from the onsets of the corresponding trials’ go signals (the white arrow, 1.5 s). These regressors were convolved with a double-gamma hemodynamic response function to be included in a general linear model (GLM) along with their temporal derivatives using FSL’s FEAT (version 5.98; Woolrich et al., 2001). Inter-trial intervals contributed to the task baseline ^32^. We used Go Fail events and fMRIPrep confound timeseries (see *MRI data preprocessing* in the supplement) as nuisance regressors. The hallmark inhibitory control contrast, Stop Success>Stop Fail, represented inhibitory control brain activity in each of the two runs.

First-level contrast estimates were entered into fixed effects models to yield subject-level statistical maps at baseline and follow-up. We first identified clusters of significant impairment in iHUD vs. HC at baseline via FSL FLAME 1&2 (FMRIB Local Analysis of Mixed Effects) with cluster-based thresholding (Z>3.1, corrected to *p*<.05; identical methods as reported in ^8^). This first step was conducted to inspect the effects we previously reported, this time in a smaller sample (the subjects who were scanned twice). Next, we inputted baseline and follow-up subject-level maps into a repeated-measures ANOVA with group (between-subjects) and session (within subjects) as fixed factors, using FSL’s non-parametric permutation testing tool, *randomise*, with threshold-free cluster enhancement (5000 permutations)—a more appropriate tool for estimating longitudinal effects compared to FLAME (https://fsl.fmrib.ox.ac.uk/fsl/fslwiki/GLM). Here we restricted the search for recovery effects via small volume correction to the clusters reflecting the baseline impairments in iHUD as compared to HC. Post-hoc t-tests inspected directionality of significant effects.

In similar repeated-measures ANOVAs, we examined ΔSSRT and Δ*d’* as covariates to detect whether increases in behavioral performance is correlated with increases in inhibitory control brain activity, corrected for familywise error (α=.05/2=.025). We further inspected the potential influence of select demographic and neuropsychological measures that showed group and/or session differences. Within iHUD, we inspected baseline heroin use-related measures and commonly used drugs that showed group differences on the time-dependent change in BOLD (see supplement). We also report cross-sectional correlational analyses in the supplement for completeness (e.g., baseline behavioral measures as correlated with baseline brain activity) but refer readers to our previous larger N study that included this sample ^8^ for their discussion.

Across all analyses, values that were three standard deviations ± the mean were identified as outliers and the affected tests are reported with and without their inclusion. Correlational analyses including these outliers were supplemented with robust regression for completeness.

## Results

### Participants

The groups were comparable in age, sex, race, nonverbal IQ, handedness, years of regular alcohol use, and years of alcohol use to intoxication (all *p*s>.201). Significant differences were noted in years of education, verbal IQ (HC>iHUD), depression symptoms, anxiety symptoms, nicotine dependence, and years of regular cannabis use (iHUD>HC); all *p*s<.002, see Table 1. Among the Table 1 variables collected at both time points, there were no significant differences between sessions in either group for depression (iHUD *p*=.398, HC *p*=.767) or anxiety symptoms (iHUD *p*=.520, HC *p*=.470). Within iHUD, cue-induced drug craving decreased at follow-up (follow-up minus baseline rating= -0.34±0.65, range= -2.06-0.72, *p*=.016). All but four iHUD remained abstinent between the two time points; on average, abstinence increased at follow-up by 65±82 days (3 missing). Cue-induced drug craving and abstinence length (at each session or their Δ) did not significantly contribute to the results below, as detailed in the supplement (which also includes the null effects of heroin use-related measures including self-reported methadone dosage at baseline).

### Behavioral Results

Despite the main effect of group for Go accuracy [HC>iHUD *F*(1,96)=11.33, *p*=.001; no session main effect: *F*(1,96)=0.04, *p*=.832, or group*session interaction: *F*(1,96)=0.13, *p*=.719] and Go RT [iHUD>HC *F*(1,96)=4.63, *p*=.034; no session main effect: *F*(1,96)=0.32, *p*=.570, or group*session interaction: *F*(1,96)<0.01, *p*=.989], all participants met the recommended performance thresholds (i.e., mean go accuracy ≥60%, mean stop accuracy ≥25% and ≤ 75%, and SSRT > 0) ^27^. There were no main effects or interactions in Stop accuracy or SSD (all *p*s>.253). See Table 2 for cross-sectional analyses where, as previously reported in a larger sample ^8^, iHUD exhibited significantly lower *d’* than HC at baseline. Indeed a significant main effect of group was evident in *d’* [HC>iHUD *F*(1,96)=4.61, *p*=.034], with no main effect of session [*F*(1,96)=0.02, *p*=.887)] or group*session interaction [*F*(1,96)=0.71, *p*=.402)]. The SSRT yielded no significant main effects of group [*F*(1,96)=2.37, *p*=.127)], session [*F*(1,96)<0.01, *p*=.953)], or group*session interaction [*F*(1,96)=1.49, *p*=.225)]. These results were not affected by education, verbal IQ, depression and anxiety, nicotine dependence, years of regular cannabis use, baseline heroin use-related measures, changes in cue-induced drug craving, or days of abstinence between sessions (see the supplement for details).

**Table 2.** Behavioral performance.

|  | Baseline |  |  | Follow-up |  |  | Group*Session |
| --- | --- | --- | --- | --- | --- | --- | --- |
|  | HC | iHUD | sig. test<sup>a</sup> | HC | iHUD | sig. test<sup>a</sup> | sig. test |
| Stop accuracy | 0.48<br>(0.04) | 0.47<br>(0.06) | $t(43.7)=0.68$,<br>$p=.497$ | 0.47<br>(0.03) | 0.49<br>(0.08) | $t(33.9)=0.96$,<br>$p=.343$ | $F(1,96)=1.32$,<br>$p=.253$ |
| Go accuracy<sup>b</sup> | 0.98<br>(0.02) | 0.96<br>(0.05) | $t(34.6)=1.95$,<br>$p=.060$ | 0.98<br>(0.03) | 0.95<br>(0.03) | $t(46.6)=3.10$,<br>$p=.003^*$ | $F(1,96)=0.13$,<br>$p=.719$ |
| SSRT | 280<br>(38.2) | 284<br>(59.9) | $t(42.8)=0.25$,<br>$p=.806$ | 266<br>(43.3) | 296<br>(69.0) | $t(42.5)=1.86$,<br>$p=.069$ | $F(1,96)=1.49$,<br>$p=.225$ |
| SSD | 272<br>(87.7) | 304<br>(142) | $t(42.2)=0.96$,<br>$p=.343$ | 296<br>(104) | 312<br>(156) | $t(43.8)=0.44$,<br>$p=.660$ | $F(1,96)=0.09$,<br>$p=.764$ |
| Go RT | 574<br>(78.1) | 625<br>(125) | $t(42.3)=1.74$,<br>$p=.089$ | 588<br>(86.8) | 639<br>(162) | $t(38.9)=1.42$,<br>$p=.164$ | $F(1,96)<0.01$,<br>$p=.989$ |
| $d'$ | 0.62<br>(0.04) | 0.58<br>(0.08) | $t(38.7)=2.37$,<br>$p=.023^*$ | 0.61<br>(0.05) | 0.59<br>(0.10) | $t(37.93)=0.87$,<br>$p=.387$ | $F(1,96)=0.71$,<br>$p=.402$ |

HC: healthy controls; iHUD: individuals with heroin use disorder; SSRT: stop-signal response time; SSD: stop-signal delay.
<sup>a</sup> Significant group differences (corrected for familywise error in two primary outcomes of interest: SSRT and $d'$ , $\alpha=.05/2=.025$ ) are flagged with an asterisk.
<sup>b</sup> Go accuracy was calculated as go responses divided by the total of correct and incorrect go responses.

### BOLD-fMRI Results

#### Cross-sectional analyses of baseline and follow-up fMRI-BOLD activity

At baseline, the whole-brain analysis of inhibitory control (Stop Success>Stop Fail) across all participants revealed significant engagement in the classical cognitive control-related nodes including the bilateral aPFC, right dlPFC, ventromedial PFC/orbitofrontal cortex, among others. When inspected between groups, iHUD displayed significantly lower Stop Success>Stop Fail signaling compared to HC in the right aPFC and right dlPFC among other regions (see eFigure 1, Supplement). No region showed significantly higher activity in iHUD compared to HC (see eTable 1 in the Supplement for details on all clusters).

At follow-up, the whole-brain analyses of inhibitory control across all participants revealed activations in the bilateral precentral gyrus among others (see eTable 2 in the Supplement for details on all clusters). No significant group differences in inhibitory control brain activity emerged at follow-up.

#### Increase of task-related brain hypoactivity following treatment

The repeated-measures ANOVA with group and session as fixed factors revealed a main effect of group in the right dlPFC and aPFC among others (see Table 3), driven by iHUD hypoactivations during inhibitory control. We found no main effect of session, but a significant group*session interaction in the right aPFC and other regions that previously indicated hypoactivity at baseline inhibitory control in the iHUD vs. HC. Post-hoc tests interrogating the direction of effects revealed a significant increase from baseline to follow-up in the right aPFC, dlPFC, among others in the iHUD (Table 3, Figure 1). The single outlier dlPFC BOLD data point in the iHUD did not affect results (the mixed model group * session interaction remained significant at *p*<.001). ΔSSRT did not correlate with increases in inhibitory control brain activity across all participants or in either group. Increases in *d’* were associated with increases in right aPFC activity following treatment in iHUD (Table 3, Figure 2), and when tested across all participants, but not within the HC. One Δ*d’* data point in the iHUD was identified as an outlier, and its exclusion reduced this correlation to a trend level when accounting for the two behavioral measures (*R*^2^=0.17, *p*=.038, α=.025). Nevertheless, a robust regression including the outliers supported the significant effect when assuming a normal t-distribution (*t*=4.04, β=0.0004, standard error=0.0001, p<.001). None of the BOLD results (baseline or change from baseline to follow-up) were significantly affected by education, verbal IQ, depression or anxiety symptoms, nicotine dependence, years of regular cannabis use, baseline heroin use-related measures, changes in cue-induced drug craving, or days of abstinence between sessions (see supplement for details).

**Table 3.**
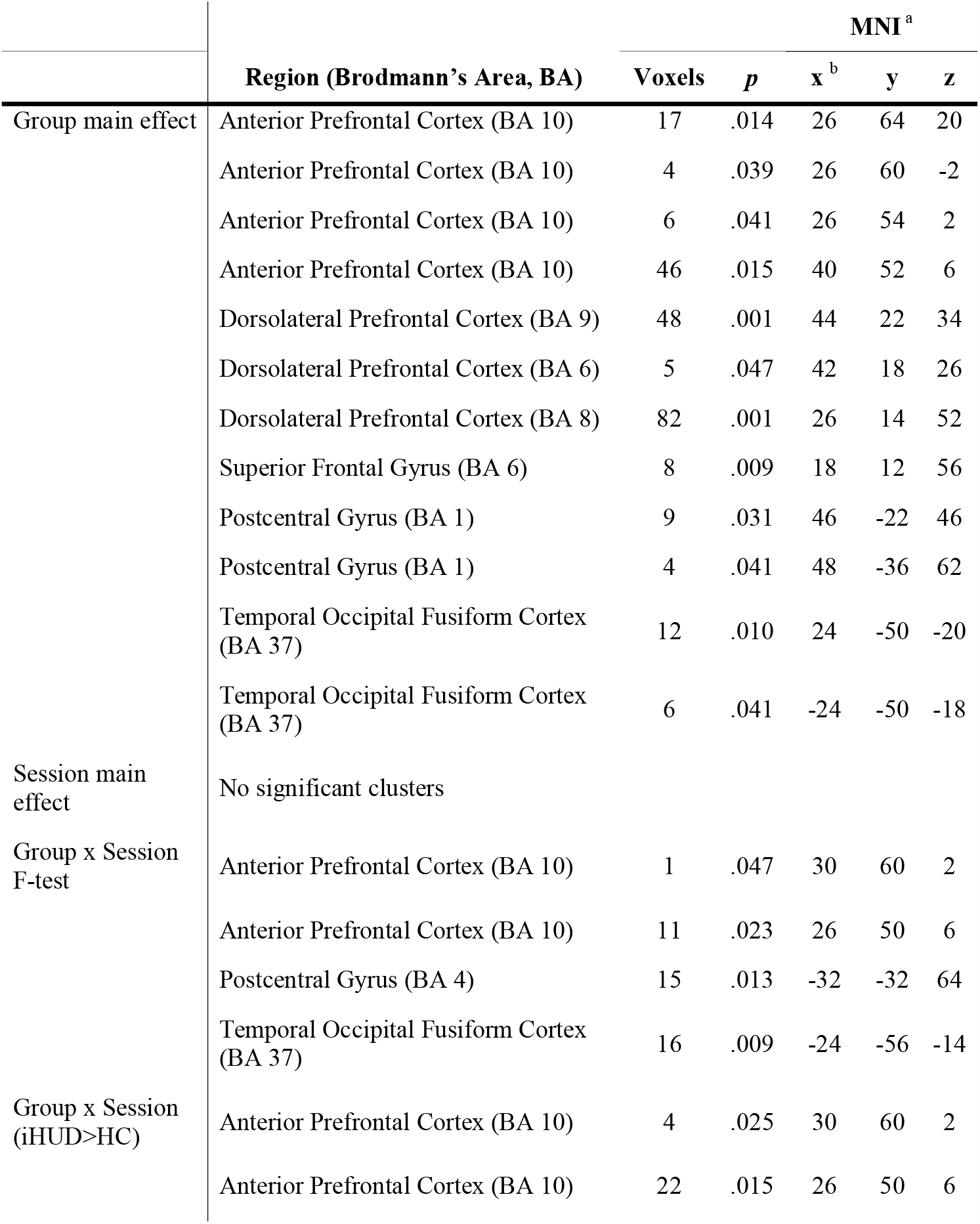
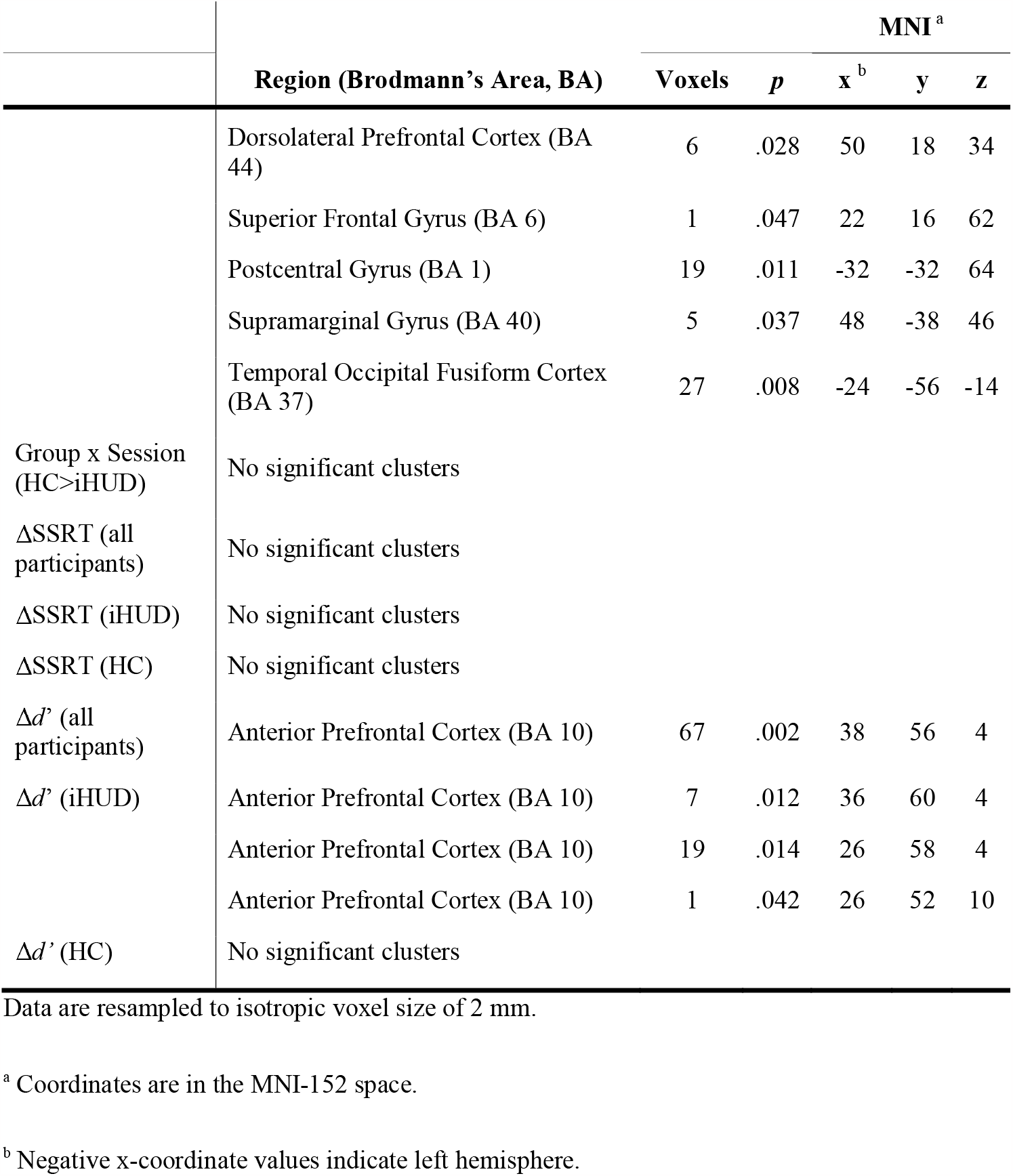
Task-related fMRI-BOLD recovery.

**Figure 1.**
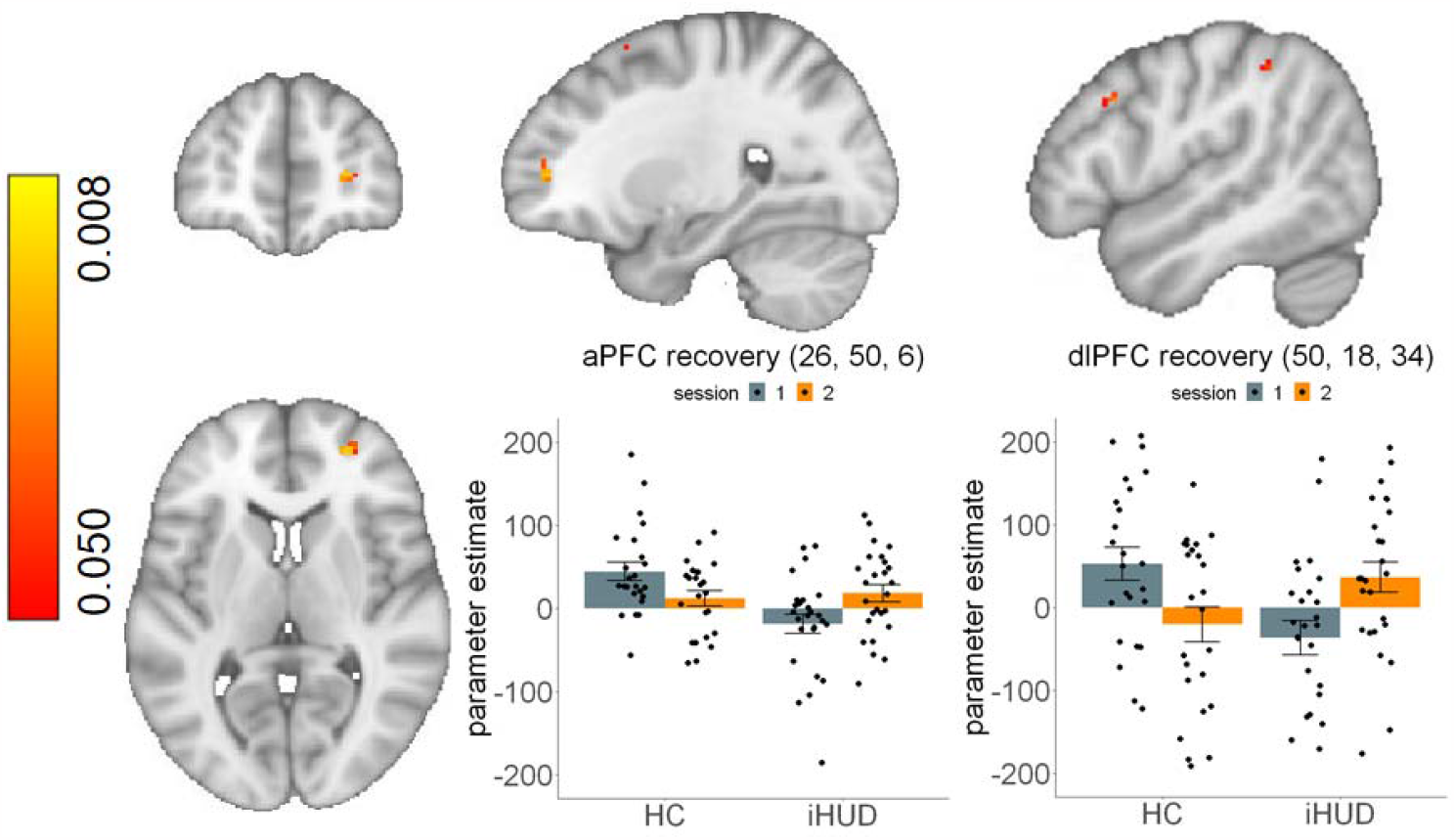
Inhibitory control brain activity increases from baseline to follow-up in individuals with heroin use disorder compared to healthy controls. Right anterior prefrontal cortex (aPFC; left plot) and right dorsolateral PFC (dlPFC; right plot) activity during successful versus failed stops showed significant increases from baseline to follow-up in individuals with heroin use disorder (iHUD) compared to healthy controls (HC). The clusters are identified by the post-hoc interrogation of the significant group (iHUD, HC) * session (baseline, follow-up) interaction (iHUD>HC, follow-up>baseline). Bar plots indicate parameter estimates from the voxel with the lowest p-value in each cluster. Swarm plots indicate individual data points. Error bars denote standard error of the mean. For visualization purposes, figure excludes single outlier data point in the iHUD that did not affect results (group * session interaction in mixed model remained significant at *p*<.001); see eFigure 2 in the Supplement for its inclusion. Coordinates are in the MNI-152 space.

**Figure 2.**
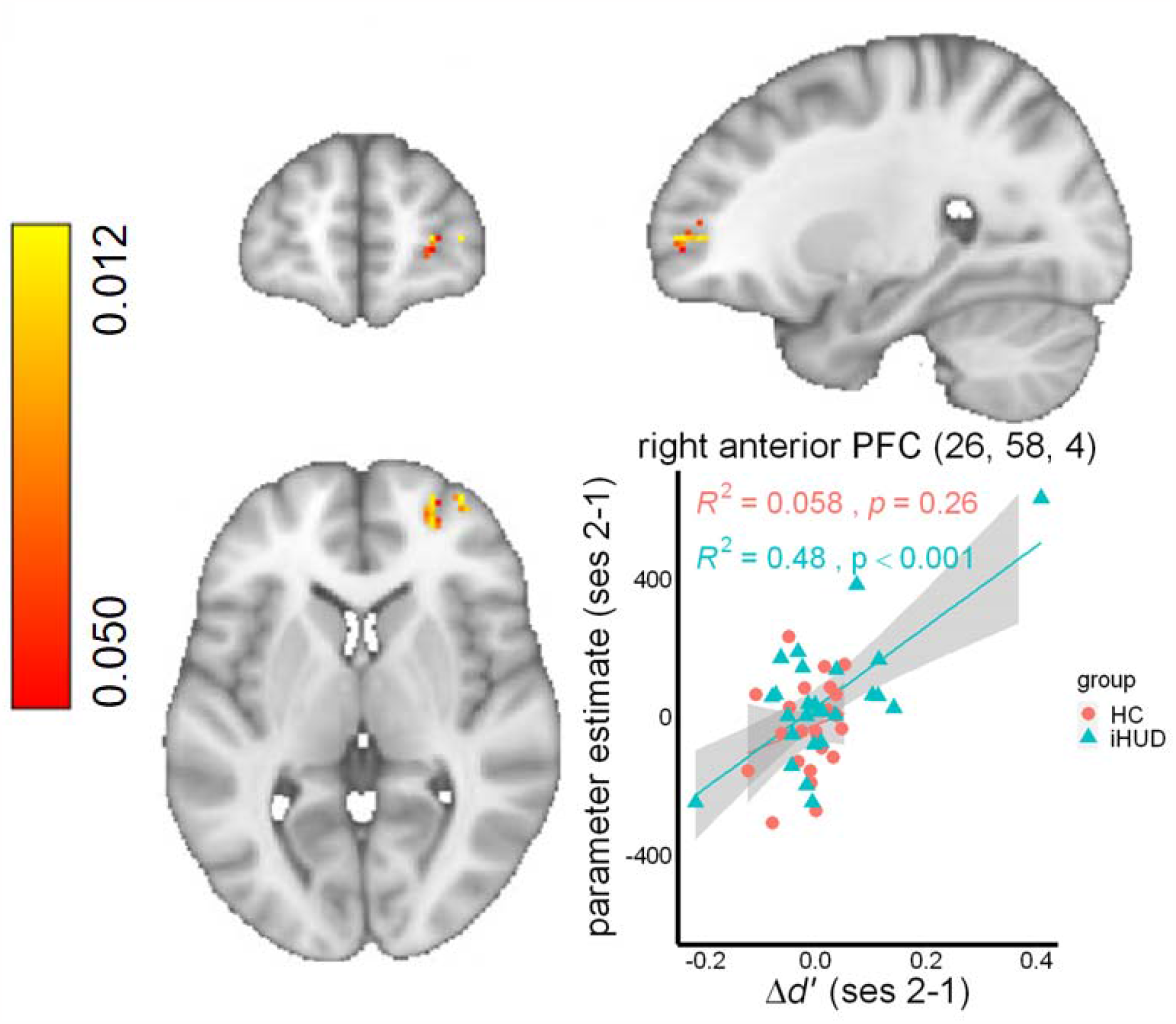
Individual differences in target detection sensitivity and inhibitory control brain activity increases. A significant relationship was evident between improvements in target detection sensitivity (Δ*d’*) and increases in right anterior prefrontal cortex (aPFC) activity during successful compared to failed stops in individuals with heroin use disorder (iHUD). One Δ*d’* data point in the iHUD was identified as an outlier, and its exclusion reduced this correlation to a trend level when accounting for two behavioral measures (including SSRT, *R*^2^=0.17, *p*=.038, α=.025). Nevertheless, a robust regression including the outliers supported the significant effect when assuming a normal t-distribution (*t*=4.04, β=0.0004, standard error=0.0001, p<.001). Coordinates are in the MNI-152 space. HC: healthy controls, ses: session.

## Discussion

For the first time in heroin (or any drug) addiction, results revealed inhibitory control brain activity enhancement after an average of 15 weeks of inpatient treatment. At baseline, compared to HC, these iHUD showed worse target detection sensitivity (yet comparable stopping speed) and lower aPFC and dlPFC activity during inhibitory control on the SST. Despite comparable performance, these PFC decrements were no longer detectible at follow-up. Importantly, these gains in inhibitory control PFC signaling were most evident in the iHUD who showed improvements in target detection sensitivity, suggesting that this BOLD signal increase may indicate a neural recovery that is related to improvement in task-relevant cognitive control processes.

This suggestion for a functional recovery of the cognitive control network (e.g., aPFC, dlPFC) during SST performance in medication-assisted inpatients with HUD extends previous longitudinal reports of general functional PFC normalizations (e.g., in cue-reactivity, decision-making, and during rest) with treatment and/or abstinence in drug addiction (see ^9^ for a review). To our knowledge, there is only one other longitudinal report in iHUD that approximated inhibitory control: after a year of methadone treatment, inpatient iHUD exhibited IFG signal increase during response selection in a Go/No-Go task ^16^. These results are consistent with a cross-sectional report where the IFG signal was higher with long vs. short-term abstinence in cocaine addiction (in the absence of Go/No-Go performance differences) ^33^. A longitudinal study of 14 medication-assisted iHUD (abstinent for three months) found decreased aPFC resting-state functional connectivity ^34^—a task-negative state that is thought to be anti-correlated with task-positive brain activity ^35^. In our study, despite comparable performance between sessions as consistent with prior research ^11,13,15,16,36^, individual differences in target detection sensitivity improvements (a behavioral measure of inhibitory control) were associated with aPFC signal increases in the iHUD compared to the HC. This neurobehavioral pattern reported here for the first time suggests a pattern of recovery that is detectable even after a short-term treatment in continuously abstinent iHUD. Given the aPFC’s role in managing task rules/goals ^37–39^, and the dlPFC’s top-down modulation of executive control ^40,41^ (see ^42,43^ for its neuromodulation improving performance in drug addiction), our results underscore the importance of individual differences in behavioral improvements as a marker of neural recovery that may inform treatment success in heroin addiction.

Several mechanisms may underlie the potential recovery of PFC function during inhibitory control in iHUD. For instance, structural reorganization of the PFC, noted to be essential for drug addiction recovery ^44^, may be related to inhibitory control functional changes. Specifically, the dlPFC BOLD signal increase is consistent with increases in gray matter volume in this region, reported with abstinence of approximately a month in alcohol ^45,46^ and six months in cocaine use disorders (in the aPFC) ^47^. Another candidate mechanism may be neuroendocrinological changes over the course of treatment. Neuropeptide Y, commonly associated with homeostatic and regulatory functions (e.g., food intake, sleep) ^48,49^, is dysregulated in neuropsychiatric conditions including substance use disorders (e.g., alcohol) ^50^. In heroin addiction, the lower neuropeptide Y levels in iHUD at acute withdrawal (3 days) approach the level of HC after a month of abstinence. Preclinical evidence suggests that a low dose administration of a selective neuropeptide Y antagonist accompanies quicker SSRT and enhanced dorsal frontal cortex-evoked striatal inhibition, potentially underlying the executive control improvements ^51^, which remains to be further tested to inform chronic heroin use and cessation. Together, increased PFC activity may be indicative of time-dependent reorganization of cognitive control processes in iHUD; more targeted and translational approaches are needed to precisely identify the neurobiological mechanisms of addiction treatment efficacy.

Our results should be interpreted in light of several limitations. First, inferences about sex- and treatment-seeking status-related contributions warrant a larger sample size with more women and a non-treatment seeking group. Relatedly, our study does not allow for the decoupling between participation in the medication-assisted treatment (in addition to the randomized group therapy) and length of abstinence (and the associated reduction in cue-induced drug craving). Such an effort may require including iHUD who are not abstinent and/or not in treatment; although note lack of significant effects on our results of both abstinence length and craving. While available baseline methadone dosage data yielded no significant correlations with outcomes of interest, we cannot rule out the potential contributions of dosage changes over the course of treatment, or type of medication (e.g., methadone vs. buprenorphine). Finally, while the variables showing group differences (years of education, verbal IQ, depression, anxiety, nicotine dependence, and years of regular cannabis use) did not correlate with outcomes of interest and thus do not explain the current results, samples more closely matched in demographics and smoking status are needed to further evaluate their potential contributions to results.

To the extent of our knowledge, this study is the first to provide empirical evidence for a time-dependent inhibitory control PFC recovery in drug addiction. Our results align with abstinence-mediated changes to a related cognitive function, response selection, previously reported in iHUD ^16^ whereby stable behavioral performance and enhanced neural activity were observed with at least three months’ follow-up compared to baseline. Including a HC group, we can rule out extraneous factors’ contributions to these results (e.g., general time and practice effects). Importantly, we identified individual differences in target detection sensitivity time-dependent changes as pertinent to the parallel neural changes, suggesting that within-subject behavioral improvements may be sensitive measures of inhibitory control PFC functional recovery. Overall, our findings suggest that aPFC and dlPFC functions could be amenable to targeted interventions (e.g., modulation of PFC activity and PFC-mediated processes through behavioral therapy, pharmacotherapy, or brain stimulation) to promote self-control, core to drug addiction symptomology, en route to recovery in iHUD.

## Data Availability

All data produced in the present study are available upon reasonable request to the authors.

## Author Contributions

The corresponding author, Rita Z. Goldstein, had full access to all of the data in the study and take responsibility for the integrity of the data and the accuracy of the data analysis.

### Concept and design

Ahmet O. Ceceli, Nelly Alia-Klein, Rita Z. Goldstein

### Acquisition, analysis, or interpretation of data

Ahmet O. Ceceli, Yuefeng Huang, Pierre-Olivier Gaudreault, Natalie E. McClain, Sarah G. King, Greg Kronberg, Amelia Brackett, Gabriela N. Hoberman, John H. Gray, Eric L. Garland, Nelly Alia-Klein, Rita Z. Goldstein

### Drafting of the manuscript

Ahmet O. Ceceli, Rita Z. Goldstein

### Critical revision of the manuscript for important intellectual content

Ahmet O. Ceceli, Yuefeng Huang, Pierre-Olivier Gaudreault, Eric L. Garland, Rita Z. Goldstein

### Statistical analysis

Ahmet O. Ceceli, Natalie E. McClain, Sarah G. King, Rita Z. Goldstein

### Administrative, technical, or material support

Ahmet O. Ceceli, Yuefeng Huang, Pierre-Olivier Gaudreault, Natalie E. McClain, Sarah G. King, Greg Kronberg, Amelia Brackett, Gabriela N. Hoberman, John H. Gray, Eric L. Garland, Nelly Alia-Klein, Rita Z. Goldstein

### Supervision

Nelly Alia-Klein, Rita Z. Goldstein

