## Supplement for "Recovery of inhibitory control prefrontal cortex function in inpatients with heroin use disorder: a 15-week longitudinal fMRI study"

**Supplementary Materials**

**eTable 1:** Task-related fMRI-BOLD activity at baseline.

**eTable 2:** Task-related fMRI-BOLD activity at follow-up.

**eFigure 1:** Inhibitory control brain activity differences between groups at baseline.

**eFigure 2:** Inhibitory control brain activity increases from baseline to follow-up in individuals with heroin use disorder compared to healthy controls, including outlier.

**Exclusion criteria**

Exclusion criteria for all participants were the following: 1) DSM-5 diagnosis for schizophrenia or neurodevelopmental disorder; 2) Head trauma with loss of consciousness longer than 30 min; 3) History of central nervous system disease; 4) Cardiovascular, metabolic, endocrinological, oncological, autoimmune, and infectious diseases including Hepatitis B and C or HIV/AIDS; 5) Metal implants or other MR contraindications (including pregnancy); 6) Court mandated treatment. We did not exclude for DSM-5 diagnosis of a drug use disorder other than opiates as long as heroin was the primary drug of choice/reason for treatment-seeking since iHUD commonly use alcohol, amphetamines, benzodiazepines, other sedatives, cocaine, and marijuana in addition to heroin. Exclusion criteria for the HC were the same, except history of any drug use disorder was prohibitive.

**Diagnostic details in individuals with heroin use disorder**

Comorbidities in iHUD included major depressive disorder (n=5), cocaine use disorder (n=4), post-traumatic stress disorder (n=3), sedative use disorder (n=3), cannabis use disorder (n=2), alcohol use disorder (n=1), generalized anxiety disorder (n=1), meth/amphetamine use disorder (n=1), obsessive-compulsive disorder (n=1), and panic disorder (n=1).

**Treatment-specific details**

Study participants were randomized into Mindfulness Oriented Recovery Enhancement and support group therapy sessions, both in addition to medication-assisted treatment. The former group sessions involved mindfulness-based self-awareness and emotion regulation training (e.g., cognitive reappraisal of negative and savoring of positive thoughts/contexts). These strategies were delivered with the goal of diminishing drug cue-reactivity while enhancing natural reward processing and cognitive control over craving and drug-seeking behaviors (see ^12^ for therapy details). The iHUD were instructed to individually partake in daily 15-min practice sessions guided by audio instructions to supplement the group sessions. The support group included structured, therapist-guided and addiction-related psychoeducation, emotional expression, and discussions, also supplemented with 15-min daily, independent journaling exercises about addiction-related topics.

**MRI data acquisition and preprocessing**

Anatomical T1-weighted images were obtained using the following parameters: 3D MPRAGE sequence with 256 × 256 × 179 mm^3^ FOV, 0.8 mm isotropic resolution, TR/TE/TI=2400/2.07/1000 msec, 8° flip angle with binomial (1, −1) fat saturation, 240 Hz/pixel bandwidth, 7.6 msec echo spacing, and in-plane acceleration (GRAPPA) factor of 2, approximate acquisition time of 7 min. The blood-oxygen-level-dependent (BOLD) fMRI responses were measured as a function of time using T2*-weighted single-shot multiband accelerated (factor of 7) gradient-echo echo-planar image (EPI) sequence [TE/TR=35/1000 ms, 2.1 isotropic mm resolution, 70 axial slices without gaps for whole-brain coverage (147 mm), FOV 206 × 181 mm, matrix size 96 × 84, 60°-flip angle (approximately Ernst angle), blipped CAIPIRINHA phase-encoding shift=FOV/3, 1860 kHz/Pixel bandwidth with ramp sampling, echo spacing 0.68 ms, and echo train length 84 ms]. The SST was administered over two, approximately 6 min 30 s, functional runs. The 1.5-hour scan session included additional structural ^1,2^ and functional ^3^ procedures reported elsewhere.

Raw BOLD-fMRI data were first converted to NIFTI and preprocessed using the fMRIprep pipeline version 20.2.1 ^4^ Structural images were intensity-normalized and skull-stripped using ANTS ^5^. These images were spatially normalized to the ICBM 152 Nonlinear Asymmetrical template using ANTS via nonlinear registration ^6,7^. Brain tissue segmentation was carried out using FSL’s FAST ^8^ to derive white matter, gray matter, and cerebrospinal fluid estimates. Functional data were corrected for motion artifacts using FSL’s MCFLIRT and for susceptibility distortion using spin-echo field maps acquired in opposing phase encoding directions via AFNI’s 3dQwarp ^9,10^. Motion- and distortion-corrected images were co-registered to the participant’s structural images using boundary-based registration with 9 degrees of freedom via FSL’s FLIRT ^7,11^. These correction, transformation, and registration steps were integrated into a single step transformation workflow using ANTS, resampled to isotropic voxels of 2 mm. The following confounds were extracted from fMRIPrep as time-series for each BOLD scan run: six translation and rotation parameters (x, y, and z for each) as motion regressors, global cerebrospinal fluid and white matter components, and cosine regressors for high-pass filtering (128 sec cutoff) to ignore low-frequency drift related to scanner and physiological noise. The preprocessed data were spatially smoothed using a Gaussian kernel (5 mm full-width at half maximum) to improve signal to noise ratio. The groups were comparable in motion at baseline (mean framewise displacement in iHUD=0.255 mm; in HC=0.209 mm, *t*(48.0)=1.46, *p*=.150) but not at follow-up (iHUD>HC, iHUD=0.278 mm; in HC=0.211 mm, *t*(45.5)=2.02, *p*=.050). As there were no significant BOLD results at follow-up, we checked for correlations between framewise displacement (motion at follow-up and Δmotion) and ΔBOLD. While Δmotion did not correlate with ΔBOLD (*r*=.245, *p*=.087), motion at follow-up positively correlated with ΔBOLD (*r*=.354, *p*=.012); however, controlling for it as a covariate in linear mixed models did not substantially affect ΔBOLD results of interest (which remained significant at *p*<.001).

**Controlling for potential confounds**

To account for the contribution of other potentially explanatory variables, we first compared iHUD and HC in age, sex, race, education, verbal IQ, non-verbal IQ, handedness, depression symptoms, anxiety symptoms, cigarette smoking status and severity of nicotine dependence, years of regular cannabis use, years of regular alcohol use, and years of alcohol use to intoxication (measures collected at baseline). We applied Welch’s two-sample t-tests, chi-square tests, or Fisher’s exact tests where appropriate, corrected for familywise error (α=.05/13=.004; cigarette smoking status was excluded given the almost parallel distribution matching group identity). Those variables showing significant group differences were tested for their potential correlation with ΔSSRT, Δ*d*’, and ΔBOLD (as well as the baseline and follow-up behavioral measures separately). If also indicating significant correlations, these variables (that showed significant group differences at baseline) served as covariates in the linear mixed models testing the time-dependent changes in our measures of interest. Within iHUD, we similarly tested these behavioral and BOLD outcomes of interest for correlations with select baseline heroin use severity measures in Table 1 [lifetime heroin use, days since last heroin use, heroin craving, withdrawal symptoms, and severity of dependence, and baseline medication dosage (corrected for familywise error: α=.05/6=.008). Heroin use in past 30 days was not inspected as inpatient iHUD largely reported no use in the past month]. Lastly, we also inspected our outcome measures for potential correlations with the variables that significantly differed between sessions in iHUD: cue-induced craving and abstinence length (at each session and their Δ), corrected for familywise error (α=.05/2=.025).

At baseline, while showing significant group differences, education, verbal IQ, depression symptoms, anxiety symptoms, nicotine dependence, and years of regular cannabis use showed no significant correlations with our dependent measures (α=.004): SSRT all *p*s >.486 in iHUD, *p*s > .103 in HC; *d’* all *p*s > .200 in iHUD, *p*s > .247 in HC; peak BOLD: *p*=.140 in iHUD, *p*=.160 in HC. There were also no significant correlations with these variables at follow-up: SSRT all *p*s > .098 in iHUD, *p*s > .060 in HC; *d’* all *p*s > .239 in iHUD, *p*s > .011 in HC, when corrected for familywise error. Correlations with peak BOLD at follow-up were not inspected as there were no significant group differences in brain activity. With the variables showing significant group differences listed above, we also found no correlations with ΔSSRT (all *p*s > .661 in iHUD, *p*s > .070 in HC), Δ*d’* (all *p*s > .132 in iHUD, *p*s > .039 in HC), or peak ΔBOLD (all *p*s > .112 in iHUD, *p*s > .188 in HC).

Within iHUD, we found no significant correlations (α=.008) between select heroin use severity measures and baseline SSRT (*p*s>.200), *d’* (*p*s>170), or peak BOLD (*p*s>.220) with the exception of a correlation between higher severity of dependence and lower peak aPFC activity at baseline (*p*=.004). Controlling for the severity of dependence as a covariate in linear mixed models did not substantially affect the lower baseline aPFC activity or time-dependent aPFC results of interest (these measures remained significant at *p*<.001). Similarly, no heroin use severity measures significantly correlated with SSRT (*p*s>.020) or *d’* (*p*s>.128) at follow-up. These variables also did not correlate with ΔSSRT (*p*=.055), Δ*d*’ (*p*=.077), or ΔBOLD (*p*=.217). Furthermore, there were no significant correlations (α=.025) between cue induced craving or abstinence length (at each session or Δ) and baseline SSRT (*ps*>.044), *d*’ (*ps>*.591), or peak BOLD activity (*ps>*.079). No correlations with SSRT (*p*s>.091) or *d*’ (*ps*>.128) were evident at follow-up. These within iHUD variables also did not correlate with ΔSSRT (*p*=.556), Δ*d*’ (*p*=.226), or ΔBOLD (*p*=.191).

**Cross-sectional correlational analyses of baseline and follow-up fMRI-BOLD activity with task performance**

At baseline, SSRT did not significantly correlate with brain activity across all participants or within iHUD, while in HC, quicker SSRT was associated with higher superior parietal lobule activity. Across all participants, higher *d’* was associated with higher bilateral aPFC and right posterior cingulate activity. While no significant clusters emerged within iHUD, in HC higher *d’* was associated with higher left dlPFC activity among others (see eTable 1 for details on all clusters).

At follow-up, across all participants, quicker SSRT was associated with lower dorsomedial PFC, paracingulate, and precuneus activity, among other regions. While no significant correlations were evident in HC, within iHUD, quicker SSRT was associated with lower activity in the posterior regions such as the precuneus and angular gyrus. Across all participants, higher *d’* was associated with higher right aPFC and right orbitofrontal cortex activity, among others. Higher *d’* correlated with higher right postcentral gyrus activity within HC, and regions including the right aPFC within iHUD (see eTable 2 for details on all clusters).

**Inhibitory control brain activity differences between groups at baseline**

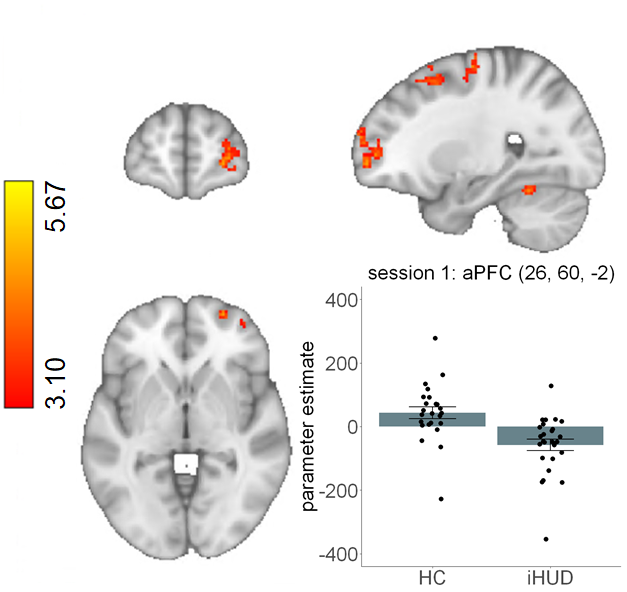

**eFigure 1. Inhibitory control brain activity differences between groups at baseline.** Individuals with heroin use disorder (iHUD), as compared to healthy controls (HC), exhibited significantly lower right anterior prefrontal cortex (aPFC) activity during successful versus failed stops. Significant results were detected using a cluster defining threshold of Z>3.1, corrected to *p*<.05. Bar plot indicates parameter estimates from the voxel with the peak Z score in each cluster. Color bar represents Z values. Swarm plots indicate individual data points. Error bars denote standard error of the mean. No data points were three standard deviations above or below the mean. Coordinates are in the MNI-152 space.

**Inhibitory control brain activity increases in iHUD vs. HC including outlier**

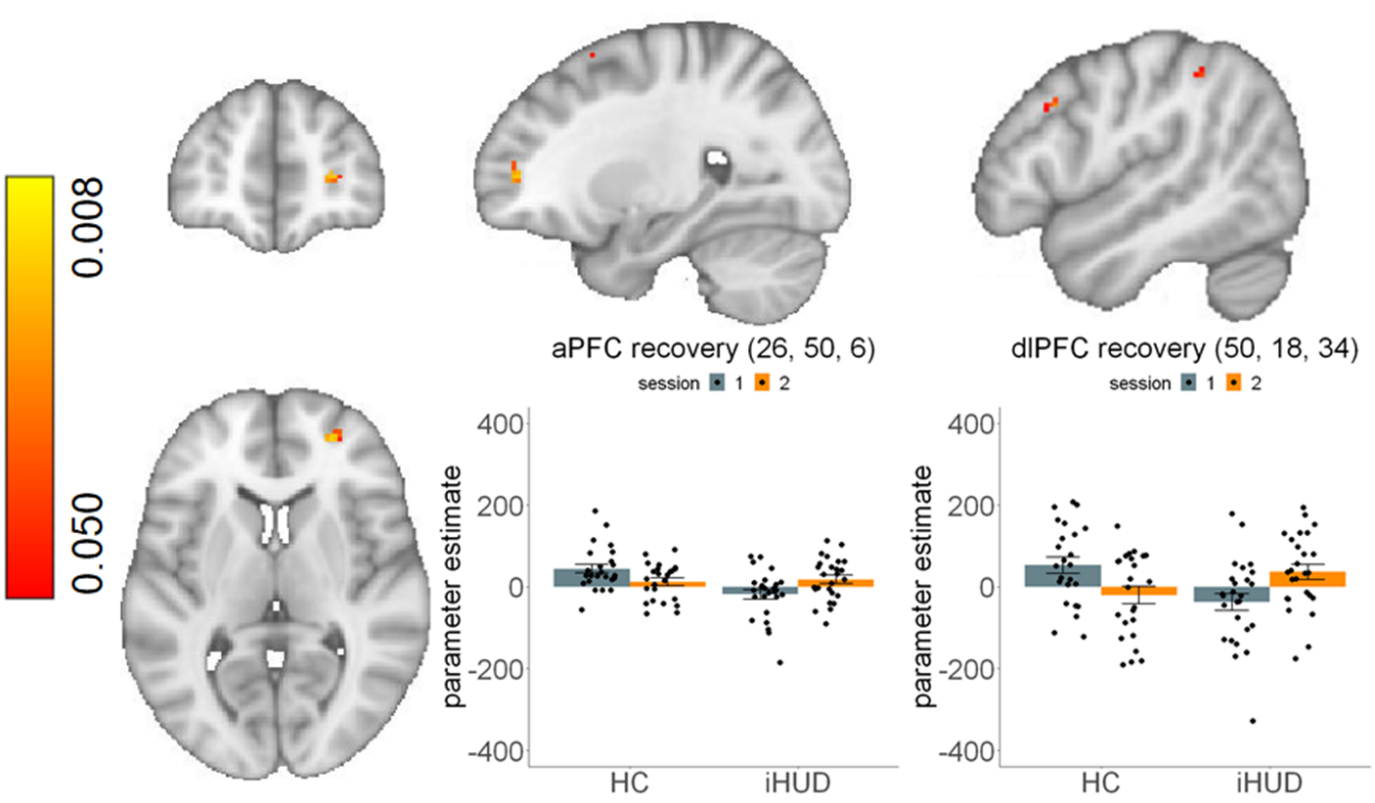

**eFigure 2. Inhibitory control brain activity increases from baseline to follow-up in individuals with heroin use disorder compared to healthy controls, including outlier.** Right anterior prefrontal cortex (aPFC; left plot) and right dorsolateral PFC (dlPFC; right plot) activity during successful versus failed stops showed significant increases from baseline to follow-up in individuals with heroin use disorder (iHUD) compared to healthy controls (HC). The clusters are identified by the post-hoc interrogation of the significant group (iHUD, HC) * session (baseline, follow-up) interaction (iHUD>HC, follow-up>baseline). Bar plots indicate parameter estimates from the voxel with the lowest p-value in each cluster. Color bar represents p-values derived from non-parametric repeated measures testing and threshold-free cluster enhancement. Swarm plots indicate individual data points. Error bars denote standard error of the mean. Figure includes single outlier data point in the iHUD that did not affect results (group * session interaction in mixed model remained significant at *p*<.001). Coordinates are in the MNI-152 space.

**Supplementary Tables**

eTable 1. Task-related fMRI-BOLD activity at baseline.

|  |  |  |  | **MNI** ^a^ | | |
| --- | --- | --- | --- | --- | --- | --- |
|  | **Region (Brodmann’s Area)** | **Voxels** | **Z** | **x** ^b^ | **y** | **z** |
| All participants | Anterior Prefrontal Cortex (BA 10) | 92 | 4.21 | 32 | 62 | -6 |
|  | Anterior Prefrontal Cortex (BA 10) | 259 | 5.19 | -36 | 58 | 4 |
|  | Orbitofrontal Cortex (BA 11) | 93 | 4.26 | 0 | 46 | -16 |
|  | Dorsolateral Prefrontal Cortex (BA 8) | 94 | 4.62 | 38 | 26 | 52 |
|  | Putamen | 74 | 4.47 | 16 | 10 | -8 |
|  | Dorsolateral Prefrontal Cortex (BA 6) | 87 | 3.9 | 30 | 0 | 50 |
|  | Lateral Occipital Cortex (BA 7) | 259 | 5.52 | 22 | -70 | 58 |
|  | Occipital Fusiform Gyrus (BA 19) | 815 | 5.6 | -34 | -82 | -14 |
|  | Occipital Fusiform Gyrus (BA 19) | 1135 | 5.69 | 30 | -84 | -12 |
|  | Lateral Occipital Cortex (BA 19) | 393 | 5.8 | -32 | -88 | 18 |
| HC>iHUD | Anterior Prefrontal Cortex (BA 10) | 269 | 4.69 | 26 | 60 | -2 |
|  | Anterior Prefrontal Cortex (BA 9) | 265 | 5.06 | 38 | 38 | 40 |
|  | Dorsolateral Prefrontal Cortex (BA 8) | 157 | 4.58 | 28 | 12 | 54 |
|  | Precentral Gyrus (BA 6) | 69 | 4.49 | 26 | -12 | 64 |
|  | Postcentral Gyrus (BA 1) | 145 | 5.01 | 46 | -24 | 48 |
|  | Postcentral Gyrus (BA 1) | 85 | 5.67 | -30 | -34 | 60 |
|  | Temporal Occipital Fusiform Cortex (BA 37) | 59 | 4.67 | 24 | -50 | -20 |
|  | Cerebellum, lobule VI | 92 | 4.22 | -24 | -64 | -22 |
| iHUD>HC | No significant clusters |  |  |  |  |  |
| SSRT, all participants | No significant clusters |  |  |  |  |  |
| SSRT, iHUD | No significant clusters |  |  |  |  |  |
| SSRT, HC | Superior Parietal Lobule (BA 7) | 65 | -4.26 | 16 | -54 | 62 |
| *d*’, all participants | Anterior Prefrontal Cortex (BA 10) | 90 | 4.45 | -20 | 58 | 16 |
|  | Anterior Prefrontal Cortex (BA 9) | 117 | 4.45 | 26 | 44 | 32 |
|  | Cingulate Gyrus, posterior division (BA 23) | 130 | 4.5 | 8 | -42 | 38 |
| *d*’, iHUD | No significant clusters |  |  |  |  |  |
| *d*’, HC | Dorsolateral Prefrontal Cortex (BA 5) | 61 | 4.27 | -54 | 12 | 38 |
|  | Temporal Occipital Fusiform Cortex (BA 37) | 70 | 4.02 | 24 | -50 | -20 |
|  | Cerebellum, lobule VI | 132 | 4.25 | -34 | -56 | -26 |
|  | Cerebellum, lobule VIIb | 57 | 4.34 | 28 | -68 | -46 |

Data are resampled to isotropic voxel size of 2 mm.

^a^ Coordinates are in the MNI-152 space.

^b^ Negative x-coordinate values indicate left hemisphere.

Supplementary Table 2. Task-related fMRI-BOLD activity at follow-up.

|  |  |  |  | **MNI** ^a^ | | |
| --- | --- | --- | --- | --- | --- | --- |
|  | **Region (Brodmann’s Area)** | **Voxels** | **Z** | **x** ^b^ | **y** | **z** |
| All participants | Putamen | 243 | 4.67 | 28 | 8 | -4 |
|  | Amygdala | 89 | 4.34 | 30 | -4 | -18 |
|  | Precentral Gyrus (BA 6) | 50 | 4.75 | 36 | -4 | 52 |
|  | Precentral Gyrus (BA 6) | 63 | 4.39 | -26 | -6 | 56 |
|  | Superior Temporal Gyrus (BA 22) | 105 | 4.62 | -58 | -10 | -2 |
|  | Temporal Occipital Fusiform Cortex (BA 37) | 96 | 4.69 | -36 | -50 | -20 |
|  | Lateral Occipital Cortex (BA 37) | 1465 | 5.95 | 44 | -62 | -12 |
|  | Lateral Occipital Cortex (BA 7) | 120 | 4.4 | 26 | -62 | 54 |
|  | Angular Gyrus (BA 39) | 125 | 4.8 | -36 | -70 | 50 |
|  | Lateral Occipital Cortex (BA 19) | 316 | 4.86 | -46 | -82 | -8 |
|  | Occipital Pole (BA 18) | 66 | 5.78 | -20 | -96 | 14 |
| HC>iHUD | No significant clusters |  |  |  |  |  |
| iHUD>HC | No significant clusters |  |  |  |  |  |
| SSRT, all participants | Paracingulate Gyrus (BA 32) | 55 | 4.01 | -6 | 44 | -6 |
|  | Superior Frontal Gyrus (BA 8) | 49 | 3.95 | 2 | 40 | 50 |
|  | Middle Temporal Gyrus (BA 21) | 52 | 3.84 | -56 | 0 | -16 |
|  | Superior Temporal Gyrus (BA 21) | 207 | 4.97 | -64 | -34 | 6 |
|  | Precuneous Cortex (BA 31) | 64 | 4.26 | -6 | -58 | 38 |
|  | Lateral Occipital Cortex (BA 39) | 182 | 4.46 | -48 | -62 | 30 |
| SSRT, iHUD | Superior Temporal Gyrus (BA 22) | 48 | 4.16 | 58 | -10 | -4 |
|  | Middle Temporal Gyrus (BA 21) | 46 | 3.88 | -56 | -12 | -14 |
|  | Precuneus Cortex (BA 31) | 46 | 4.2 | -2 | -56 | 38 |
|  | Angular Gyrus (BA 39) | 87 | 4.13 | -42 | -58 | 26 |
| SSRT, HC | No significant clusters |  |  |  |  |  |
| *d*’, all participants | Anterior Prefrontal Cortex (BA 10) | 87 | 4.17 | 26 | 58 | 18 |
|  | Orbitofrontal Cortex (BA 47) | 50 | 4.43 | 52 | 24 | -10 |
|  | Temporal Pole (BA 20) | 60 | 4.24 | 48 | 8 | -38 |
|  | Temporal Pole (BA 34) | 56 | 4.65 | -26 | 4 | -18 |
|  | Postcentral Gyrus (BA 1) | 120 | 5.11 | 68 | -12 | 14 |
|  | Parietal Operculum Cortex (BA 40) | 50 | 4.11 | 48 | -22 | 14 |
|  | Hippocampus | 190 | 4.77 | 30 | -28 | -10 |
|  | Hippocampus | 56 | 4.27 | -22 | -30 | -8 |
|  | Postcentral Gyrus (BA 1) | 51 | 4.28 | 26 | -38 | 68 |
|  | Cingulate Gyrus, posterior division (BA 30) | 85 | 4.4 | -4 | -44 | 4 |
|  | Precuneous Cortex (BA 31) | 102 | 4.89 | -4 | -46 | 50 |
|  | Middle Cerebellar Peduncle | 49 | 3.97 | 20 | -46 | -36 |
|  | Cerebellum, lobule VI | 155 | 3.85 | -18 | -54 | -30 |
|  | Lingual Gyrus (BA 18) | 113 | 4.53 | -6 | -90 | -16 |
| *d*’, iHUD | Anterior Prefrontal Cortex (BA 10) | 50 | 4.28 | 24 | 56 | 16 |
|  | Middle Temporal Gyrus (BA 20) | 154 | 4.66 | 54 | -2 | -32 |
|  | Postcentral Gyrus (BA 1) | 105 | 5.06 | 68 | -12 | 14 |
|  | Hippocampus | 58 | 4.38 | 22 | -16 | -16 |
|  | Hippocampus | 75 | 4.15 | -22 | -30 | -8 |
|  | Cingulate Gyrus, posterior division (BA 31) | 86 | 4.21 | 6 | -34 | 48 |
|  | Superior Temporal Gyrus (BA 22) | 55 | 4.22 | 66 | -36 | 4 |
|  | Precuneous Cortex (BA 31) | 76 | 4.52 | -6 | -40 | 50 |
|  | Cerebellum, lobule I-IV | 144 | 4.39 | 0 | -42 | -28 |
|  | Planum Temporale (BA 41) | 76 | -4.26 | -38 | -42 | 8 |
|  | Temporal Occipital Fusiform Cortex (BA 19) | 72 | 4.36 | 26 | -52 | -8 |
|  | Cerebellum, lobule V | 63 | 4.37 | 0 | -62 | -12 |
|  | Lingual Gyrus (BA 18) | 52 | 4.58 | 6 | -86 | -20 |
| *d*’, HC | Postcentral Gyrus (BA 4) | 195 | 4.5 | 56 | -6 | 34 |

Data are resampled to isotropic voxel size of 2 mm.

^a^ Coordinates are in the MNI-152 space.

^b^ Negative x-coordinate values indicate left hemisphere.
